# Temporal Aspects of the Association between Exposure to the World Trade Center Disaster and Risk of Skin Melanoma

**DOI:** 10.1101/2021.03.10.21253261

**Authors:** Paolo Boffetta, David G. Goldfarb, Rachel Zeig-Owens, Dana Kristjansson, Jiehui Li, Robert M. Brackbill, Mark R. Farfel, James E. Cone, Janette Yung, Amy R. Kahn, Baozhen Qiao, Maria J. Schymura, Mayris P. Webber, David J. Prezant, Christopher R. Dasaro, Andrew C. Todd, Charles B. Hall

**Author notes:** **Corresponding Author**: Paolo Boffetta, MD, MPH, Stony Brook Cancer Center, Stony Brook University, Lauterbur Dr., Stony Brook, NY 11794 USA.

## Abstract

Rescue/recovery workers who responded to the World Trade Center (WTC) attacks on 9/11/2001 were exposed to known/suspected carcinogens. Studies have identified an increased risk of skin melanoma in this population, but the temporal aspects of the association have not been investigated. A total of 44,540 non-Hispanic White workers from the *WTC Combined Rescue/Recovery Cohort* were observed between 3/12/2002 and 12/31/2015. Cancer data were obtained via linkages with 13 state registries. Poisson regression was used to estimate hazard ratios (HR) and 95% confidence intervals (CI), using the New York State population as reference; change points in the HRs were estimated using profile likelihood. We observed 247 incident cases of skin melanoma. No increase in incidence was detected between 2002 and 2004. Beginning in 2005, the HR was 1.34 (95% CI 1.18-1.52). A dose-response relationship was observed according to time worked on the WTC effort. Risk of melanoma among non-Hispanic White WTC rescue/recovery workers was elevated, beginning in 2005. While WTC-related exposure to ultraviolet radiation or other agents might have contributed to this result, exposures other than the WTC effort and enhanced medical surveillance cannot be discounted. Our results support the continued surveillance of this population for melanoma.

## INTRODUCTION

The terrorist attacks of the World Trade Center (WTC) on 9/11/2001, in addition to their immediate lethal consequences among people present at the site and rescue/recovery workers, caused widespread, persistent exposure to toxic substances (Landrigan et al., 2004). Carcinogens known to be present in relatively high quantities at the WTC site included asbestos, benzene, chromium, dioxins and polychlorinated biphenyls (PCBs) (Claudio, 2001, Lioy and Georgopoulos, 2006, Lioy et al., 2002).

The incidence of malignant skin melanoma (hereafter, melanoma), has increased over the last decades in fair-skinned persons in the US and other countries (Erdmann et al., 2013). This trend has been attributed to increased exposure to natural (solar) and artificial ultraviolet (UV) radiation (Gandini et al., 2005). Other occupational and environmental agents, however, are suspected to increase the risk of melanoma as well, including polycyclic aromatic hydrocarbons (PAHs) (Sim et al., 2020), metalworking fluids (Costello et al., 2011), arsenic (Matthews et al., 2019), ionizing radiation (Fink and Bates, 2005) and PCBs (Boffetta et al., 2018).

An increased risk of melanoma has been reported in previous analyses of three cohorts of WTC-exposed rescue/recovery workers (Li et al., 2012, Moir et al., 2016, Shapiro et al., 2020). Because of the small number of observed events, the excesses did not reach statistical significance because of limited power and could not be investigated in detail. In addition, the interpretation of these early results is complicated by several factors, including differences in the definition of the exposed and the reference populations and in analytical approach to estimate cancer risk, as well as the overlap across the cohorts (Boffetta et al., 2016). Additionally, there were differences in the time periods studied between WTC exposure and cancer occurrence.

To better characterize the risk of melanoma in non-Hispanic White workers with WTC exposure, including the temporal relationship of diagnosis with exposure, we analyzed the incidence of this neoplasm in the *WTC Combined Rescue/Recovery Cohort* (Brackbill et al., 2021), which comprises workers enrolled in the three cohorts mentioned above. By examining the temporal aspects of the experience of cancer in this combined population, we aim to address the scientific question regarding the latency between an environmental carcinogen exposure and the appearance of elevated incidence of melanoma among non-Hispanic White individuals.

## RESULTS

Among 44,540 participants in the final analytic cohort, we observed 247 melanoma cases for 241 participants between 3/12/2002 and 12/31/2015, with 491,492 person-years of follow-up. During this period in the NYS reference population, there were 46,233 cases of melanoma diagnosed, with 134,922,302 person-years. Demographic characteristics for the analytic cohort are presented in Table 1. A total of 9.1% of cases occurred among female responders, whereas females comprised 12.4% of responders without melanoma (p<0.01); the mean age at diagnosis among melanoma cases was 55.9 (SD=11.5) and ranged from 28 to 88. Among melanoma cases, the median time from 9/11 to diagnosis was 9.9 years (interquartile range, 6.5, 12.1). Among cases of melanoma, 28 (11.6%) had another malignancy within the study period; this proportion was 6.3% among persons who were diagnosed with a cancer other than melanoma in the study period (n=2,785). Fourteen of the cases of melanoma (5.8%) died during the follow-up, compared with 3.1% of persons without melanoma (p<0.05).

**Table 1.** Selected characteristics of study population

| **Characteristics** | **Melanoma cases (n=241)** | **Other rescue/recovery workers<sup>a</sup> (n=44,299)** | **Total (n=44,540)** |
| --- | --- | --- | --- |
| **Age at study entry (mean, SD)** | 49.0 (11.8) | 43.3 (10.4) | 43.4 (10.4) |
| **Sex** |  |  |  |
| Male | 219 (90.9) | 38,793 (87.6) | 39,012 (87.6) |
| Female | 22 (9.1) | 5,506 (12.4) | 5,528 (12.4) |
| **Vital status by the end of follow-up (12/31/2015) (n, %)** |  |  |  |
| Deceased | 14 (5.8) | 1,380 (3.1) | 1,394 (3.1) |
| Alive | 227 (94.2) | 42,919 (96.9) | 43,146 (96.9) |
| **Cohort membership (n, %)** |  |  |  |
| FDNY<sup>b</sup> | 104 (43.2) | 13,898 (31.4) | 14,002 (31.4) |
| GRC<sup>c</sup> | 80 (33.2) | 16,820 (38.0) | 16,900 (37.9) |
| WTCHR<sup>d</sup> | 57 (23.7) | 13,581 (30.7) | 13,638 (30.6) |
| **First worked at WTC site (n, %)** |  |  |  |
| 9/11/2001 - 9/17/2001 | 187 (77.6) | 34,278 (77.4) | 34,465 (77.4) |
| 9/18/2001 – 6/30/2002 | 38 (15.8) | 7,574 (17.1) | 7,612 (17.1) |
| Not at main WTC site | 8 (3.3) | 1,853 (4.2) | 1,861 (4.2) |
| Missing | 8 (3.3) | 594 (1.3) | 602 (1.4) |
| **Worked on pile (n, %)** |  |  |  |
| Yes | 119 (49.4) | 19,873 (44.9) | 19,992 (44.9) |

Legend: FDNY (Fire Department of the city of New York); GRC (General Responder Cohort); WTCHR (World Trade Center) Health Registry; WTC (World Trade Center)
a=Rescue/recovery workers that did not have a skin melanoma diagnosis in the study period; b=includes participants followed by the WTCHR and/or the GRC; c=includes participants followed by the WTCHR; d=not followed by FDNY or GRC

Tumor site and stage among the combined cohort and NYS reference data are presented in Table 2. The combined cohort had a higher proportion of localized tumors (75.7% vs 71.4%), a lower proportion of regional tumors (5.7% vs 9.3%; p<0.05), and a similar proportion of distant tumors (4.5% vs 4.2%) when compared with NYS. Age-standardized rates were higher among WTC rescue/recovery workers for localized and regional tumors. The majority of tumors among WTC rescue/recovery workers were located on the trunk (n=112, 45.3%), followed by the upper limb/shoulder (n=45, 18.2%) and lower limb/hip (n=32, 13.0%).

**Table 2.** Selected clinical characteristics of skin melanoma cases

| **Primary Site** | **Description** | **WTC-RR n**<br>**(row %)** | **Rate** | **NYS**<br>**n (row %)** | **Rate** |
| --- | --- | --- | --- | --- | --- |
| C445 | Skin of trunk | 112 (45.3)* | 19.0 | 15,990 (33.8) | 11.2 |
| C446 | Skin of upper limb and shoulder | 45 (18.2) | 11.4 | 11,844(24.1)* | 8.2 |
| C447 | Skin of lower limb and hip | 32 (13.0) | 7.6 | 9,089 (19.3)* | 6.5 |
| C444 | Skin of scalp and neck | 22 (8.9)* | 5.3 | 3,194 (6.6) | 2.2 |
| C443 | Skin of other and unspecified parts of face | 18 (7.3) | 3.8 | 4,626 (9.0)* | 3.1 |
| C449 | Skin, NOS | 11 (4.5) | 1.4 | 2,104 (4.1) | 1.2 |
| C442 | External ear | ≤5 (1.6) | 1.0 | 1,336 (2.8) | 0.9 |
| C448 | Overlapping lesion of skin | ≤5 (0.8)* | 0.4 | 62 (0.0) | 0.0 |
| C441 | Skin of eyelid | ≤5 (0.4) | 0.1 | 186 (0.3) | 0.1 |
| **Stage** |  | **WTC-RR n**<br>**(row %)** | **Rate** | **NYS**<br>**n (row %)** | **Rate** |
|  | Localized | 187 (75.7)* | 39.6 | 34,657 (71.4) | 24.2 |
|  | Regional | 14 (5.7) | 3.3 | 4,490 (9.3)* | 3.1 |
|  | Distant | 11 (4.5) | 1.4 | 2,021 (4.2) | 1.4 |
|  | Unknown | 35 (14.2) | 5.5 | 7,346 (15.1) | 5.1 |

Legend: WTC-RR (*WTC Combined Rescue/Recovery Cohort*); NYS (New York State) reference population; Rates are age and sex standardized to the US 2000 non-Hispanic White population;
\*=Chi square p-value is less than <0.05 for all categories.

Figure 1 displays adjusted incidence rates for the study. Rates were based on piecewise exponential models with no change points and were centered at male participants aged 50-59 in the secondary models. The incidence of melanoma among the combined cohort was greater than that of NYS throughout the whole study period and increased with follow-up.

**FIGURE 1.**
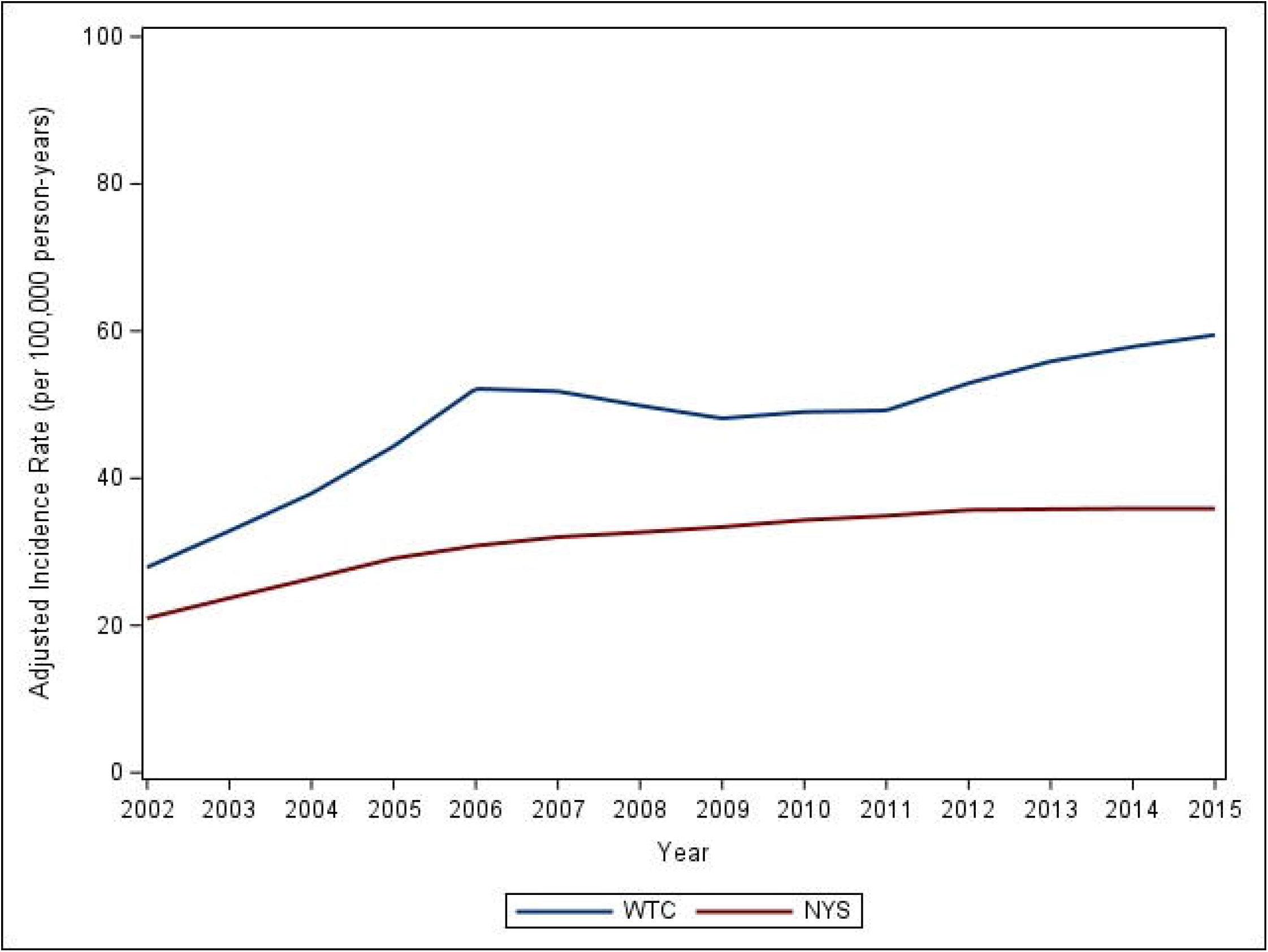
Adjusted incidence rate of melanoma of the skin in the *WTC Combined Rescue/Recovery Cohort* and in the New York State population, 2002-2015 WTC (World Trade Center); NYS (New York State) Analysis restricted to non-Hispanic Whites. Rates are adjusted for sex and age and are centered at males aged 50-59.

Table 3 shows the results of the analyses when comparing to the external NYS reference population. For our model, which did not evaluate change points, we observed an HR of 1.31 for melanoma (95% CI 1.15-1.48) for the whole study period (2002-2015). In our change point analysis, we estimated a change point in 2004, and the elevation in the HR was restricted to the period from 2005 to 2015 (HR 1.34; 95% CI 1.18-1.52). In our analysis which evaluated localized tumors only as the outcome, we observed no change points and an increased hazard compared to NYS (HR 1.36; 95% CI 1.18-1.57). When evaluating regional/distant tumors only, model estimates were largely unstable because of the small number of tumors (results not presented).

**Table 3.** Hazard ratio of skin melanoma by WTC responder status. Results of comparison with NYS rates

A. Overall
| N change points | Period of follow-up | N cases | Person-years | HR | 95% CI |
| --- | --- | --- | --- | --- | --- |
| 0 | 2002-2015 | 247 | 491,492 | 1.31 | 1.15-1.48 |
| 1 | 2002-2004 | 6 | 44,731 | 0.65 | 0.30-1.49 |
|  | 2005-2015 | 241 | 446,761 | 1.34 | 1.18-1.52 |

B. Localized
| N change points | Period of follow-up | N cases | Person-years | HR | 95% CI |
| --- | --- | --- | --- | --- | --- |
| 0 | 2002-2015 | 187 | 491,492 | 1.36 | 1.18-1.57 |

Legend: HR (Hazard Ratio); WTC (World Trade Center); NYS (New York State)
0 change point model estimates the hazard over the course of the study period, 2002-2015; 1 change point model is independent and estimates the hazard in two distinct intervals, from 2002-2004 and 2005-2015; all models are adjusted for age, sex and calendar year

The results of the internal analyses are reported in Table 4. In the analysis by time-period-of-work on the WTC effort, we identified a change point in 2009, and the HR during 2010-2015 for work between 9/11/2001 and 9/17/2001 was significantly elevated. This result contrasts with our external model above, which identified a change point in 2004. In the analysis examining the effect of working on the WTC pile, we identified a change point in 2004, but the results were limited by the small number of events in the period between 2002 and 2004. In the zero-change point model, we observed an increased hazard among those who worked on the pile compared to those who did not (HR=1.32, 95% CI 1.00, 1.74).

**Table 4.** Hazard ratio of skin melanoma by period of work at WTC site. Results of internal comparison

a. Adjusted for age, sex and calendar year
| N change points | Period of follow-up | First worked at WTC site | N cases | Person-years | HR | 95% CI |
| --- | --- | --- | --- | --- | --- | --- |
| 0 | 2002-2015 | Early | 191 | 386,865 | 1.28 | 0.89, 1.85 |
|  |  | Late | 40 | 82,820 |  | ref |
| 1 | 2002-2009 | Early | 66 | 186,643 | 0.68 | 0.42, 1.09 |
|  |  | Late | 26 | 38,536 |  | ref |
|  | 2010-2015 | Early | 125 | 200,222 | 2.41 | 1.36, 4.27 |
|  |  | Late | 14 | 44,285 |  | ref |

b. Adjusted for age, sex, calendar year and cohort membership
| N change points | Period of follow-up | First worked at WTC site | N cases | Person-years | HR | 95% CI |
| --- | --- | --- | --- | --- | --- | --- |
| 0 | 2002-2015 | Early | 191 | 386,865 | 1.03 | 0.69, 1.52 |
|  |  | Late | 40 | 82,820 |  | ref |
| 1 | 2002-2009 | Early | 66 | 186,643 | 0.54 | 0.32, 0.88 |
|  |  | Late | 26 | 38,536 |  | ref |
|  | 2010-2015 | Early | 125 | 200,222 | 1.93 | 1.07, 3.48 |
|  |  | Late | 14 | 44,285 |  | ref |

Legend: HR (Hazard Ratio); WTC (World Trade Center); CI (Confidence interval)
0 change point model estimates the hazard over the course of the study period, 2002-2015; 1 change point model is independent and estimates the hazard in two distinct intervals, from 2002-2009 and 2010-2015
Early=first self-reported working at the main WTC disaster site between 9/11/2001-9/17/2001;
Late=first self-reported working at the WTC disaster site between 9/18/2001-6/30/2002.
Melanoma cases among persons who did not self-report working at the main WTC site between 9/11/2001-6/30/2002 were excluded from all analysis (n=16 persons; 16 cancer cases).

## DISCUSSION

In the current study, we observed an increased incidence of melanoma among non-Hispanic White participants of the *WTC Combined Rescue/Recovery Cohort*. Earlier assessments among participants enrolled in the FDNY, GRC and WTCHR cohorts have independently demonstrated an elevated risk for melanoma; however, none showed a statistically significant relationship. Li and coworkers (Li et al., 2012) studied 21,218 rescue/recovery workers enrolled in the WTCHR during 2002-2003, who were followed for cancer incidence until 2009. These authors observed 24 cases of malignant melanoma (standardized incidence ratio [SIR], using the population of NYS as reference, 1.48; 95% confidence interval [CI] 0.97-2.16). Moir and colleagues (Moir et al., 2016) compared the incidence of cancer during 2001-2009 in 11,457 WTC-exposed firefighters to that of 8,220 non-WTC exposed firefighters from three urban areas of the US. They observed 40 cases of melanoma in WTC-exposed firefighters and 21 cases in non-WTC-exposed firefighters (relative risk [RR] 1.69; 95% CI 0.93-3.13). Finally, Shapiro and colleagues (Shapiro et al., 2020) investigated cancer incidence during 2002-2013 among 28,729 rescue and recovery workers who were enrolled in the GRC, using the population of the state of residence as reference. These authors observed 50 cases of melanoma (SIR 1.15; 95% CI 0.86-1.52).

The present analysis corroborates these early findings and identifies a period when the risk appears to be greatest. We observed no association between WTC exposure and melanoma cancer compared to NYS before 2005, and a 34% increased risk from 2005 until the end of the study period in 2015. However, this difference is based on a small number of cases occurring during 2002-2004 and may be due to an overrepresentation of FDNY cohort members in the early period of follow-up. In our internal analysis, which uses those who were least exposed as the referent category, we identified a change point in 2009 with an elevated risk among those present at the WTC rescue/recovery site during the first week after the attacks. The external analysis, which includes participants who were present at the WTC rescue/recovery site any time from 9/11/2001 through 6/30/2002 in the exposed category, resulted in an earlier change point of 2004.

Data on the temporal aspects of the association between environmental carcinogens and melanoma risk refer primarily to UV exposure. Studies on age at migration from areas with low ambient sunlight to areas with high ambient sunlight, and vice versa, provided strong evidence that UV exposure in childhood is a strong risk factor for melanoma occurrence and death in adults (Holman and Armstrong, 1984, Khlat et al., 1992, Mack and Floderus, 1991). Younger age at migration to sunnier areas has a stronger effect on the risk of melanoma than duration of residence in these areas. In addition, the risk of melanoma seems to be lower in the absence of significant sun exposure during childhood (Autier and Dore, 1998). These results reinforce the notion that heavy exposure to the sun during childhood would be a major factor determining the occurrence of melanoma in adult life. In this respect, the latency period of three years detected in the present analysis may reflect the carcinogenicity of exposures other than UV radiation experienced on the WTC effort, and also other phenomena such as environmental and occupational exposures that occurred outside the WTC experience, or enhanced surveillance.

The *WTC combined Rescue/Recovery cohort* includes firefighters, law enforcement and construction workers. An increased risk of melanoma has consistently been reported in previous studies of firefighters (Casjens et al., 2020) and police officers (Finkelstein, 1998, Forastiere et al., 1994, Harris et al., 2018), while results of studies of construction workers are mainly negative (Alicandro et al., 2020, Gallagher et al., 1987, Hakansson et al., 2001, Robinson et al., 1995). Workplace exposure to UV radiation has been suggested to explain these findings, since other occupational groups with opportunity of UV exposure, such as offshore workers (Stenehjem et al., 2017), airline pilots and cabin crews (Sanlorenzo et al., 2015), and agricultural workers (Kachuri et al., 2017) also experience an increased incidence of melanoma. However, the excess in melanoma risk is mainly observed among subjects with intermittent, prolonged UV exposure (Autier and Dore, 2020). These findings do not support the hypothesis of a causal role of WTC-related UV exposure that, on average, was of relatively short duration (i.e., <1-9 months). We cannot exclude a role of UV exposure experienced by cohort members outside WTC-related operations. The role of other agents that were present at the WTC site, such as PAHs and PCBs (Lioy and Georgopoulos, 2006) is also plausible, although an estimate of their contribution is complicated by the lack of exposure data.

Increased medical surveillance is an alternative explanation of our findings. An increase in the incidence of melanoma, in particular the more superficial forms of the disease following enhanced surveillance, has been described in several populations (Armstrong and Kricker, 1994, Bagley et al., 1981) and is supported by the increased rate of early-stage melanomas among rescue/recovery workers.

The main strengths of our study include greater statistical power compared to previous analyses of WTC rescue and recovery workers, leading to a detailed assessment of temporal aspects of the association and the ability to conduct internal dose-response analyses. Also, by including 13 different states in the linkages of the cohort to central cancer registries, we were able to cover 93% of the addresses of the 44,540-member study population. Earlier works reported the suggestion of an elevated risk among WTC-exposed participants; however, this is the first study that observed a significantly elevated risk of over 30% for 247 cases of melanoma. In our internal analysis, after adjusting for individual WTC cohort (which may be an important proxy for access to insurance and cancer coverage), the association was attenuated but still significant.

Limitations include the lack of quantitative data of exposure to carcinogens in WTC rescue/recovery operations, as well as the lack of information on the potential confounding effects of occupational and recreational exposures to UV and other agents outside the WTC experience. Another weakness was that we did not have adequate power to study melanoma in non-White races. Other authors have found that among non-White racial groups, particularly African Americans, the incidence of skin melanoma is lower overall, yet with a higher proportion of advanced stage tumors (Cormier et al., 2006, Cress and Holly, 1997, Wang et al., 2016). It was observed that, after controlling for stage, relative mortality is similar (Mahendraraj et al., 2017, Ward-Peterson et al., 2016), and the high incidence of advanced stage tumors may reflect poorer access to preventative care. Thus, further investigation of the role of latency in skin melanoma is needed in more ethnically diverse cohorts.

In conclusion, our analysis confirmed an increased incidence of melanoma among non-Hispanic White WTC rescue/recovery workers. Compared to the NYS population, the increase in incidence started after 2004 and persisted despite the extension of the follow-up. This result, and the result from internal analyses showing a higher risk for those who responded shortly after the attacks, are consistent with a contributory role of WTC exposure, but alternative explanations cannot be excluded. Regardless of the precise causes, the results support the continued surveillance of this population for melanoma.

## MATERIALS AND METHODS

### Study population

The study population included all eligible rescue/recovery workers from three WTC-exposed responder cohorts: the Fire Department of the City of New York (FDNY) (Webber et al., 2011), the General Responder Cohort (GRC) (Herbert et al., 2006) and the World Trade Center Health Registry (WTCHR) (Farfel et al., 2008). Rescue/recovery workers included firefighters, emergency medical service (EMS) providers, law enforcement, construction and communication workers, volunteers and cleanup workers. All adult rescue/recovery workers who were members of any of the three cohorts and provided informed consent for research or who fell under a waiver of consent approved by the IRB of the cohort institution were eligible for inclusion in the study. Additional details regarding consolidation of the *WTC Combined Rescue/Recovery Cohort* (hereafter, combined cohort), including de-duplication of subjects and data harmonization, are described elsewhere (Brackbill et al., 2021).

A total of 24,562 members of the combined cohort were excluded from the analysis for the following reasons: (i) less than 18 years of age on 9/11 (N=165), (ii) missing date of birth (N=21), (iii) enrolled in cohort after 10/1/2012 (N=4,402), and (iv) race and ethnicity other than non-Hispanic White (N=19,974). Only 7 (0.04%) of the 19,974 participants with race/ethnicity other than non-Hispanic White had melanoma. The current analysis is therefore based on 44,540 non-Hispanic, White rescue/recovery workers enrolled in the cohort before 10/1/2012 (i.e., when cancer coverage began under the James Zadroga 9/11 Health and Compensation Act) (Centers for Disease Control and Prevention, 2020) and were at least 18 years old on 9/11/2001.

### Outcome assessment

Person-time accruals began on the later of 3/12/2002 or six months after date of enrollment into a WTC rescue/recovery cohort. The date of 3/12/2002 was chosen so that prevalent cancers already developed before 9/11/2001 were not misclassified as incident cases; six months was chosen to reduce selection in the cohorts following the onset of symptoms. The follow-up period ended at the earlier of death or 12/31/2015. Incident cases of melanoma defined (using the SEER site recode table [25010]) as ICD-O-3 topography code C44, and malignant behavior code 3, were obtained by matching the combined cohort to data from the cancer registries of the following states: Arizona, California, Connecticut, Florida, Massachusetts, New Jersey, New York, North Carolina, Ohio, Pennsylvania, Texas, Virginia, and Washington. The thirteen states accounted for 93.1% of the known addresses of the 99.5% of persons with known address who were included in the study. We were missing state of residence for 113 of the 44,540 persons included in the analysis. Tumor characteristics, such as diagnosis date and stage, were provided by state cancer registries. Cases of cancers obtained from multiple registries for the same participant were identified and duplicates were excluded. For the calculations of age-at-diagnosis and person-time-at-risk, the 15^th^ of each month was used. For the few participants who were missing birth month, June was used. Death dates available for each cohort were reconciled by the New York State (NYS) cancer registry (NYSCR) to ensure precise mortality ascertainment and person-time contributions.

### Exposure measur es and demographic characteristics

The specific measures used for this study included dust exposure on 9/11 and work periods on the WTC effort (Weakley et al., 2011). The exposure measure for our primary external analysis was presence at the WTC site or WTC rescue/recovery work at other sites such as the Fresh Kills Landfill, at any time from 9/11/2001 to 6/30/2002, the time when the WTC Manhattan site closed. We used two additional exposure variables: (i) work on the WTC pile (yes or no); (ii) time of first arrival at the WTC disaster sites (9/11/2001-9/17/2001 or 9/18/2001-06/30/2002. Basic demographic characteristics such as sex, birth month, birth year, and race/ethnicity were used for data analysis.

### NYS Comparison Population

Incident melanoma tumors in NYS were selected as the reference for our external analysis and were obtained and organized using SEER*stat software (National Cancer Institute, 2020). Data were summarized in strata of persons and cases by age (in 5-year increments), sex and calendar year (2002 to 2015).

### Statistical Analyses

We first calculated age-standardized incidence rates by primary site and stage for the combined cohort and the NYS comparison population. Rates were standardized to the US 2000 male and female populations. The outcome for all multivariable analyses was incident melanoma. Both comparison rates and observed cancer counts included multiple primary cancers for each person. That is, persons with melanoma were counted as a case even if they had cancer before the start of follow-up for the present study (i.e., the later of 3/12/2002 or 6 months after enrollment). Persons could be counted more than once if they were diagnosed with more than one melanoma. The combined cohort data were grouped in strata of person-time and cases in the same way as the NYS comparison population.

We used piecewise, exponential survival models to estimate hazard ratios (HRs) and associated 95% confidence intervals (CIs). This approach is similar to Cox regression but allows baseline hazard to change at numerous time intervals rather than with every event; one-year time intervals were used (Jensen and Lutkebohmert, 2008). This model also allowed for incidence to be estimated in the reference group, so that the HRs have rate ratio interpretations. The HRs could vary through the follow-up period using change points.

The specific model is as follows: Let *Y*_*ik*_ be the number of incident cases of melanoma, modeled to follow a Poisson distribution given the covariates; *T*_*ik*_ is the total person-time at risk, for each particular stratum *i* and time interval *k*; *t*_*ik*_ is the time since exposure, with exposures indicated by the values of *x*_*i*_, a binary variable taking the value of 1 for the WTC-exposed cohort and 0 for the comparison NYS population; and *z*_*il*_ represents sex, calendar year and age. The *w*_*ik*_’s are dummy variables representing the one-year time intervals. β_*j*_ is the log HR, comparing incidence in the WTC-exposed cohort to the NYS reference population for time interval *j*, which is the period between change points *t*_*j-1*_ and *t*_*j*_. The change points are estimated using profile likelihood (Goodman et al., 2011, Hall et al., 2000, Hall et al., 2003, Hall et al., 2001).The *a*_*k*_’s are parameters representing the baseline hazard as a function of time that has cases-per-person-time interpretation. Equation 1 is found below and described in greater detail elsewhere (Jensen and Lutkebohmert, 2008). This primary external analysis, using NYS as a comparison, controlled for age and calendar year.

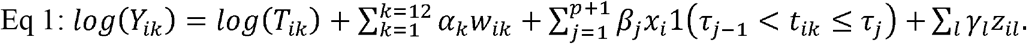

Four additional analyses were conducted. The first was an external analysis that evaluated localized tumors, separately, to address the potential for surveillance bias. The second was an internal analysis that evaluated two distinct periods of time during which participants first worked on the WTC effort (9/11/2001-9/17/2001; 9/18/2001-6/30/2002), hereafter referred to as early and late; these mutually exclusive categories were included as binary variables. The third was an internal analysis that evaluated participants that worked on the WTC pile vs those who did not. Finally, the fourth model controlled for WTC cohort membership (i.e., FDNY, GRC, and WTCHR), as well as for the other covariates used in the second analysis. We controlled for cohort membership to account for an uneven distribution of period which participants first worked at the WTC site (e.g., FDNY had the highest proportion of participants who first worked at the WTC site in the early period). For each analysis, change points were separately evaluated.

The same model equation applied to internal models as to the external model, except that the WTC variable was replaced with time-first-worked on the WTC effort. We also calculated adjusted baseline incidence rates stratified by exposure category. For this analysis, we applied a LOESS smoothing function for point estimates (Hastie and Tibshirani, 1986).

Analyses were performed using SAS 9.4 (SAS Institute Inc. Cary, NC). This study followed the Strengthening the Reporting of Observational Studies in Epidemiology (STROBE) reporting guidelines (von Elm et al., 2007) and was approved by Institutional Review Boards at Albert Einstein College of Medicine, New York City Department of Health and Mental Hygiene, the NYS Department of Health, and all thirteen cancer registries. The Icahn School of Medicine at Mount Sinai IRB ruled the research exempt.

## Data Availability

Data that support the findings of the study may be obtained from the corresponding author (PB) upon reasonable request after approval by the Steering Committee for "Incidence, Latency, and Survival of Cancer Following World Trade Center Exposure" (NIOSH Cooperative Agreement U01 OH011932) in accordance with the study's official Data Sharing Plan.

## ACKNOWLEDGEMENTS

We thank the 13 state cancer registries for carrying out record linkages: Bureau of Cancer Epidemiology, New York State Department of Health (DOH); Arizona Cancer Registry Department of Health Services (DOHS); California Cancer Registry, Department of Public Health (DPH); Connecticut Tumor Registry, Connecticut DPH; Florida Cancer Registry, Florida DOH; Massachusetts Cancer Registry, Massachusetts DPH; New Jersey State Cancer Registry, New Jersey DOH and Rutgers Cancer Institute of New Jersey; North Carolina Central Cancer Registry, State Center for Health Statistics; Ohio Cancer Incidence Surveillance System, Ohio DOH; Bureau of Health Statistics and Research, Pennsylvania DOH; Texas Cancer Registry, Texas Department of State Health Services, Virginia Cancer Registry, Virginia DOH, and Washington State Cancer Registry, Washington DOH.

Additional Acknowledgements and Disclaimers from individual State Cancer Registries: The collection of cancer incidence data used in this study was supported by the California Department of Public Health pursuant to California Health and Safety Code Section 103885; Centers for Disease Control and Prevention’s (CDC) National Program of Cancer Registries, under cooperative agreement 5NU58DP006344; the National Cancer Institute’s Surveillance, Epidemiology and End Results Program under contract HHSN261201800032I awarded to the University of California, San Francisco, contract HHSN261201800015I awarded to the University of Southern California, and contract HHSN261201800009I awarded to the Public Health Institute. The ideas and opinions expressed herein are those of the author(s) and do not necessarily reflect the opinions of the State of California, Department of Public Health, the National Cancer Institute, and the Centers for Disease Control and Prevention or their Contractors and Subcontractors.

The Connecticut Department of Public Health Human Investigations Committee approved this research project, which used data obtained from the Connecticut Department of Public Health. The Connecticut Department of Public Health does not endorse or assume any responsibility for any analyses, interpretations or conclusions based on the data. The authors assume full responsibility for all such analyses, interpretations and conclusions.

The Florida cancer incidence data used in this report were collected by the Florida Cancer Data System (FCDS), the statewide cancer registry funded by the Florida Department of Health (DOH) and the Centers for Disease Control and Preventions National Program of Cancer Registries (CDC-NPCR). The views expressed herein are solely those of the author(s) and not necessarily reflect those of the DOH or CDC-NPCR.

Cancer incidence data used in these analyses were obtained from the Ohio Cancer Incidence Surveillance System (OCISS), Ohio Department of Health (ODH), a cancer registry partially support by the National Program of Cancer Registries at the Centers for Disease Control and Prevention (CDC) through Cooperative Agreement Number NU58DP006284. Use of these data does not imply that ODH or CDC agrees or disagrees with the analyses, interpretations or conclusions in this report (or publication/presentation).

These data were supplied by the Bureau of Health Statistics & Registries, Pennsylvania Department of Health, Harrisburg, Pennsylvania. The Pennsylvania Department of Health specifically disclaims responsibility for any analyses, interpretations or conclusions.

Additional Contributions: Molly Skerker, MPH provided substantial administrative support in the initial phase of this study. She did not receive financial compensation for this work.

## AUTHOR CONTRIBUTIONS

Conceptualization: PB, DGG, RZO, JEC, MJS, CBH; Funding Acquisition: PB, CBH; Methodology: PB, DGG, CBH; Writing-Original Draft Preparation: PB, DGG, RZO, CBH; Writing-Reviewing and Editing: ALL AUTHORS

## Abbreviations

WTC: (World Trade Center)
FDNY: (Fire Department of the City of New York)
GRC: (General Responder Cohort)
WTCHR: (World Trade Center Health Registry)
NYS: (New York State)
HR: (Hazard Ratio)

## REFERENCES

Alicandro G, Bertuccio P, Sebastiani G, La Vecchia C, Frova L. Mortality among Italian male workers in the construction industry: a census-based cohort study. Eur J Public Health 2020;(30(2):247–52.

Armstrong BK, Kricker A. Cutaneous melanoma. Cancer Surv 1994;19-20:219-40.

Autier P, Dore JF. Influence of sun exposures during childhood and during adulthood on melanoma risk. EPIMEL and EORTC Melanoma Cooperative Group. European Organisation for Research and Treatment of Cancer. Int J Cancer 1998;(77(4):533–7.

Autier P, Dore JF. Ultraviolet radiation and cutaneous melanoma: a historical perspective. Melanoma Res 2020;(30(2):113–25.

Bagley FH, Cady B, Lee A, Legg MA. Changes in clinical presentation and management of malignant melanoma. Cancer 1981;(47(9):2126–34.

Boffetta P, Catalani S, Tomasi C, Pira E, Apostoli P. Occupational exposure to polychlorinated biphenyls and risk of cutaneous melanoma: a meta-analysis. Eur J Cancer Prev 2018;(27(1):62–9.

Boffetta P, Zeig-Owens R, Wallenstein S, Li J, Brackbill R, Cone J, et al. Cancer in World Trade Center responders: Findings from multiple cohorts and options for future study. Am J Ind Med 2016;(59(2):96–105.

Brackbill RK, Kahn AR, Li J, Zeig-Owens R, Goldfarb DG, Skerker M, et al. Combining Three Cohorts of WTC Rescue/Recovery Workers for Assessing Cancer Incidence and Mortality. Preprints 2021;10.20944/preprints202101.0105.v1.

Casjens S, Bruning T, Taeger D. Cancer risks of firefighters: a systematic review and meta-analysis of secular trends and region-specific differences. Int Arch Occup Environ Health 2020;(93(7):839–52.

Centers for Disease Control and Prevention. Covered Conditions −World Trade Center Health Program, https://www.cdc.gov/wtc/conditions.html; 2020 [accessed 01/01/2021.

Claudio L. Environmental aftermath. Environ Health Perspect 2001;(109(11):A528–36.

Cormier JN, Xing Y, Ding M, Lee JE, Mansfield PF, Gershenwald JE, et al. Ethnic differences among patients with cutaneous melanoma. Arch Intern Med 2006;(166(17):1907–14.

Costello S, Friesen MC, Christiani DC, Eisen EA. Metalworking fluids and malignant melanoma in autoworkers. Epidemiology 2011;(22(1):90–7.

Cress RD, Holly EA. Incidence of cutaneous melanoma among non-Hispanic whites, Hispanics, Asians, and blacks: an analysis of california cancer registry data, 1988-93. Cancer Causes Control 1997;(8(2):246–52.

Erdmann F, Lortet-Tieulent J, Schuz J, Zeeb H, Greinert R, Breitbart EW, et al. International trends in the incidence of malignant melanoma 1953-2008--are recent generations at higher or lower risk? Int J Cancer 2013;(132(2):385–400.

Farfel M, DiGrande L, Brackbill R, Prann A, Cone J, Friedman S, et al. An overview of 9/11 experiences and respiratory and mental health conditions among World Trade Center Health Registry enrollees. J Urban Health 2008;(85(6):880–909.

Fink CA, Bates MN. Melanoma and ionizing radiation: is there a causal relationship? Radiat Res 2005;(164(5):701–10.

Finkelstein MM. Cancer incidence among Ontario police officers. Am J Ind Med 1998;(34(2):157–62.

Forastiere F, Perucci CA, Di Pietro A, Miceli M, Rapiti E, Bargagli A, et al. Mortality among urban policemen in Rome. Am J Ind Med 1994;(26(6):785–98.

Gallagher RP, Elwood JM, Threlfall WJ, Spinelli JJ, Fincham S, Hill GB. Socioeconomic status, sunlight exposure, and risk of malignant melanoma: the Western Canada Melanoma Study. J Natl Cancer Inst 1987;(79(4):647–52.

Gandini S, Sera F, Cattaruzza MS, Pasquini P, Picconi O, Boyle P, et al. Meta-analysis of risk factors for cutaneous melanoma: II. Sun exposure. Eur J Cancer 2005;(41(1):45–60.

Goodman MS, Li Y, Tiwari RC. Detecting multiple change points in piecewise constant hazard functions. J Appl Stat 2011;(38(11):2523–32.

Hakansson N, Floderus B, Gustavsson P, Feychting M, Hallin N. Occupational sunlight exposure and cancer incidence among Swedish construction workers. Epidemiology 2001;(12(5):552–7.

Hall CB, Lipton RB, Sliwinski M, Stewart WF. A change point model for estimating the onset of cognitive decline in preclinical Alzheimer’s disease. Stat Med 2000;19(11-12):1555–66.

Hall CB, Ying J, Kuo L, Lipton RB. Bayesian and profile likelihood change point methods for modeling cognitive function over time. Computational Statistics & Data Analysis 2003;(42(1):91–109.

Hall CB, Ying J, Kuo L, Sliwinski M, Buschke H, Katz M, et al. Estimation of bivariate measurements having different change points, with application to cognitive ageing. Stat Med 2001;(20(24):3695–714.

Harris MA, Kirkham TL, MacLeod JS, Tjepkema M, Peters PA, Demers PA. Surveillance of cancer risks for firefighters, police, and armed forces among men in a Canadian census cohort. Am J Ind Med 2018;(61(10):815–23.

Hastie T, Tibshirani R. Generalized Additive Models. Statist Sci 1986;(1(3):297–310.

Herbert R, Moline J, Skloot G, Metzger K, Baron S, Luft B, et al. The World Trade Center disaster and the health of workers: five-year assessment of a unique medical screening program. Environ Health Perspect 2006;(114(12):1853–8.

Holman CD, Armstrong BK. Cutaneous malignant melanoma and indicators of total accumulated exposure to the sun: an analysis separating histogenetic types. J Natl Cancer Inst 1984;(73(1):75–82.

Jensen U, Lutkebohmert C. A Cox-type regression model with change-points in the covariates. Lifetime Data Anal 2008;(14(3):267–85.

Kachuri L, Harris MA, MacLeod JS, Tjepkema M, Peters PA, Demers PA. Cancer risks in a population-based study of 70,570 agricultural workers: results from the Canadian census health and Environment cohort (CanCHEC). BMC Cancer 2017;(17(1):343.

Khlat M, Vail A, Parkin M, Green A. Mortality from melanoma in migrants to Australia: variation by age at arrival and duration of stay. Am J Epidemiol 1992;(135(10):1103–13.

Landrigan PJ, Lioy PJ, Thurston G, Berkowitz G, Chen LC, Chillrud SN, et al. Health and environmental consequences of the world trade center disaster. Environ Health Perspect 2004;(112(6):731–9.

Li J, Cone JE, Kahn AR, Brackbill RM, Farfel MR, Greene CM, et al. Association between World Trade Center exposure and excess cancer risk. JAMA 2012;(308(23):2479–88.

Lioy PJ, Georgopoulos P. The anatomy of the exposures that occurred around the World Trade Center site: 9/11 and beyond. Ann N Y Acad Sci 20061076:54–79.

Lioy PJ, Weisel CP, Millette JR, Eisenreich S, Vallero D, Offenberg J, et al. Characterization of the dust/smoke aerosol that settled east of the World Trade Center (WTC) in lower Manhattan after the collapse of the WTC 11 September 2001. Environ Health Perspect 2002;(110(7):703–14.

Mack TM, Floderus B. Malignant melanoma risk by nativity, place of residence at diagnosis, and age at migration. Cancer Causes Control 1991;(2(6):401–11.

Mahendraraj K, Sidhu K, Lau CSM, McRoy GJ, Chamberlain RS, Smith FO. Malignant Melanoma in African-Americans: A Population-Based Clinical Outcomes Study Involving 1106 African-American Patients from the Surveillance, Epidemiology, and End Result (SEER) Database (1988-2011). Medicine (Baltimore) 2017;(96(15):e6258.

Matthews NH, Fitch K, Li WQ, Morris JS, Christiani DC, Qureshi AA, et al. Exposure to Trace Elements and Risk of Skin Cancer: A Systematic Review of Epidemiologic Studies. Cancer Epidemiol Biomarkers Prev 2019;(28(1):3–21.

Moir W, Zeig-Owens R, Daniels RD, Hall CB, Webber MP, Jaber N, et al. Post-9/11 cancer incidence in World Trade Center-exposed New York City firefighters as compared to a pooled cohort of firefighters from San Francisco, Chicago and Philadelphia (9/11/2001-2009). Am J Ind Med 2016;(59(9):722–30.

National Cancer Institute D, Surveillance Research Program. Surveillance, Epidemiology, and End Results (SEER) Program (www.seer.cancer.gov) SEER*Stat Database: Incidence -SEER Research Data, 9 Registries, Nov 2019 Sub (1975-2017) - Linked To County Attributes - Time Dependent (1990-2017) Income/Rurality, 1969-2017 Counties, released April 2020, based on the November 2019 submission. 2020.

Robinson C, Stern F, Halperin W, Venable H, Petersen M, Frazier T, et al. Assessment of mortality in the construction industry in the United States, 1984-1986. Am J Ind Med 1995;(28(1):49–70.

Sanlorenzo M, Wehner MR, Linos E, Kornak J, Kainz W, Posch C, et al. The risk of melanoma in airline pilots and cabin crew: a meta-analysis. JAMA Dermatol 2015;(151(1):51–8.

Shapiro MZ, Wallenstein SR, Dasaro CR, Lucchini RG, Sacks HS, Teitelbaum SL, et al. Cancer in General Responders Participating in World Trade Center Health Programs, 2003-2013. JNCI Cancer Spectr 2020;(4(1):pkz090.

Sim MR, Tan SH, Kelly S, Nixon RL. Malignant Neoplasms of the Skin. In: Anttila S, Boffetta P, editors. Occupational Cancers, 2nd Ed. Cham, Switzerland: Springer; 2020. p. pp. 401–16.

Stenehjem JS, Robsahm TE, Bratveit M, Samuelsen SO, Kirkeleit J, Grimsrud TK. Ultraviolet radiation and skin cancer risk in offshore workers. Occup Med (Lond) 2017;(67(7):569–73.

von Elm E, Altman DG, Egger M, Pocock SJ, Gotzsche PC, Vandenbroucke JP, et al. The Strengthening the Reporting of Observational Studies in Epidemiology (STROBE) statement: guidelines for reporting observational studies. Ann Intern Med 2007;(147(8):573–7.

Wang Y, Zhao Y, Ma S. Racial differences in six major subtypes of melanoma: descriptive epidemiology. BMC Cancer 2016;16:691.

Ward-Peterson M, Acuna JM, Alkhalifah MK, Nasiri AM, Al-Akeel ES, Alkhaldi TM, et al. Association Between Race/Ethnicity and Survival of Melanoma Patients in the United States Over 3 Decades: A Secondary Analysis of SEER Data. Medicine (Baltimore) 2016;(95(17):e3315.

Weakley J, Maslow C, Group. Ww. Defining common categories of exposure among four cohorts of rescue/recovery workers who responded to the World Trade Center disaster. Council of State and Territorial Epidemiologists (CSTE) Annual Conference. Pittsburgh, PA2011.

Webber MP, Glaser MS, Weakley J, Soo J, Ye F, Zeig-Owens R, et al. Physician-diagnosed respiratory conditions and mental health symptoms 7-9 years following the World Trade Center disaster. Am J Ind Med 2011;(54(9):661–71.

